# Functional genomics implicates natural killer cells in the pathogenesis of ankylosing spondylitis

**DOI:** 10.1101/2023.09.21.23295912

**Authors:** Marcos Chiñas, Daniela Fernandez-Salinas, Vitor R. C. Aguiar, Victor E. Nieto-Caballero, Micah Lefton, Peter A. Nigrovic, Joerg Ermann, Maria Gutierrez-Arcelus

## Abstract

**Objective:** Multiple lines of evidence indicate that ankylosing spondylitis (AS) is a lymphocyte-driven disease. However, which lymphocyte populations are critical in AS pathogenesis is not known. In this study, we aimed to identify the key cell types mediating the genetic risk in AS using an unbiased functional genomics approach.

**Methods:** We integrated genome-wide association study (GWAS) data with epigenomic and transcriptomic datasets of human immune cells. To quantify enrichment of cell type-specific open chromatin or gene expression in AS risk loci, we used three published methods that have successfully identified relevant cell types in other diseases. We performed co-localization analyses between GWAS risk loci and genetic variants associated with gene expression (eQTL) to find putative target genes.

**Results:** Natural killer (NK) cell-specific open chromatin regions are significantly enriched in heritability for AS, compared to other immune cell types such as T cells, B cells, and monocytes. This finding was consistent between two AS GWAS. Using RNA-seq data, we validated that genes in AS risk loci are enriched in NK cell-specific gene expression. Using the human Space-Time Gut Cell Atlas, we also found significant upregulation of AS-associated genes predominantly in NK cells. Co-localization analysis revealed four AS risk loci affecting regulation of candidate target genes in NK cells: two known loci, *ERAP1 and TNFRSF1A*, and two under-studied loci, *ENTR1* (aka *SDCCAG3*) and *B3GNT2*.

**Conclusion:** Our findings suggest that NK cells may play a crucial role in AS development and highlight four putative target genes for functional follow-up in NK cells.

## Introduction

Axial spondyloarthritis (axSpA) is a chronic inflammatory rheumatic disease characterized by inflammation of the spine and sacroiliac joints, with a proportion of patients also presenting with arthritis in peripheral joints, uveitis, psoriasis or inflammatory bowel disease (1). Historically, most genetic and pathogenetic studies in axSpA have been carried out in ankylosing spondylitis (AS), a severe and well-characterized subtype of axSpA. The heritability of AS is high, with estimates ranging between 40-90% (2). HLA-B27 is the major risk allele for AS (OR = 21.4) (3). Additionally, genome wide-association studies (GWAS) have revealed >100 non-MHC risk loci for AS, most of them implicating non-coding variants (4–8).

Many immune cell-types have been associated with axSpA (9,10). However, which ones are “driver” cell types actively contributing to the pathogenesis of the disease, as opposed to “bystanders” that become involved as a consequence of the disease, remains unclear. Studies leveraging genetic risk variants and their overlap with epigenomic and transcriptomic features variably suggested CD8+ T cells, CD4+ T cells, NK (natural killer) cells, monocytes, and gastrointestinal cells as potential mediators of AS genetic risk (10–14). However, these studies did not apply the new functional genomics datasets generated from human cells or the latest methodologies designed to integrate functional genomics with GWAS data. This new generation of methods takes advantage of the full range of SNPs examined in a GWAS (not just those surpassing the genome-wide significance threshold) and robustly control for genomic and linkage disequilibrium biases (15–17).

For several immune-mediated diseases, these integrative functional genomics methods have successfully identified specific cell types as drivers of disease development. For example, for rheumatoid arthritis (RA), multiple studies have found a significant enrichment of genetic risk in open or active chromatin regions (marking regulatory elements) specific for T cells (11,18–20). Both mouse and human studies corroborate the role of T cells as central players in RA pathogenesis (21–23). Similarly, for systemic lupus erythematosus (SLE), studies have identified an enrichment of B cell-specific putative regulatory elements and gene expression in SLE risk loci (19,20,24,25) consistent with the well-established role of B cells in SLE pathogenesis (26,27). Hence, there is precedence that the integration of GWAS with functional genomics datasets can identify cellular drivers in inflammatory diseases with complex pathogenesis.

Here we sought to investigate which immune cell populations could be drivers of AS development. We integrated GWAS summary statistics from two different AS cohorts with epigenomic and transcriptomic datasets of human leukocytes from peripheral blood and tissue using established methods that control for biases in genomic enrichment analyses. Our results bring forward NK cells as potential key drivers in the pathogenesis of AS.

## Results

In order to assess which immune cell types might be mediating the genetic susceptibility to AS, we first utilized a dataset of open chromatin profiles of immune cell subsets from peripheral blood of four healthy subjects (19) (**Fig. 1A**). Sorted cell subsets were analyzed using ATAC-seq with or without prior *in vitro* stimulation. For our study, we grouped the cells analyzed by Calderon et al. into 7 main immune cell types: T cells, B cells, NK cells, plasma cells, dendritic cells (DCs), plasmacytoid DCs, and monocytes. We identified cell type-specific open chromatin regions and assessed whether these were significantly enriched in AS genetic risk. We used the LDSC-SEG method (15) to quantify enrichment of partitioned heritability in each of these cell type-specific annotations (conceptual scheme in **Fig. 1B**, data in **Fig. 1C**) compared to baseline and control annotations, while taking into account the effects of linkage disequilibrium. We excluded the MHC region from our analyses given the unusually high linkage disequilibrium in this region and the fact that genetic associations with this locus are mostly driven by coding variants of the HLA-B gene. Using the Immunochip association study summary statistics from the International Genetics of Ankylosing Spondylitis Consortium (IGAS) (7), we found that NK cell-specific open chromatin regions were significantly enriched in genetic risk for AS (P = 0.026), while this was not the case for the other six immune cell types (**Fig. 1D**).

**Figure 1.**
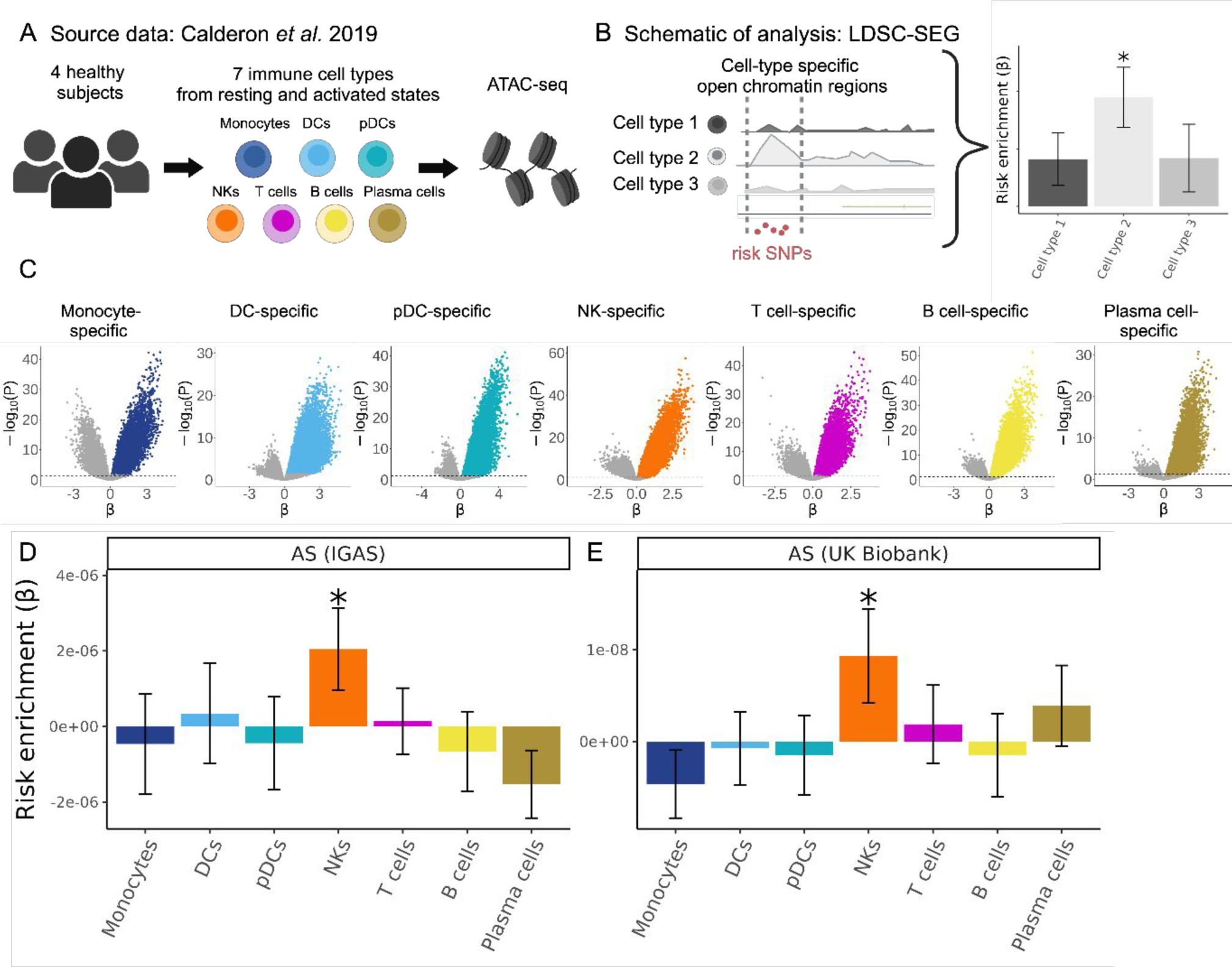
Human NK cell-specific open chromatin regions are enriched in AS genetic risk. (**A**) Cartoon depicting the Calderon *et al.* study design. Peripheral blood cells from 4 healthy subjects were sorted into immune cell populations that we grouped *in silico* into seven cell types (see Methods). Assay for Transposase-Accessible Chromatin using sequencing (ATAC-seq) was performed with and without prior *in vitro* activation. (**B**) Graphical representation of LDSC-SEG analysis: identification of cell type-specific annotations (in our case open chromatin regions), followed by the integration with GWAS summary statistics to obtain a risk enrichment coefficient β and P-value. (**C**) Volcano plots showing results of differential accessibility analyses for each cell type compared to the other cell types. Colored dots indicate open chromatin peaks in the top decile of the t-statistic for each cell type, which were used for LDSC-SEG analysis. (**D-E**) Bar graphs displaying the AS genetic risk enrichment coefficient β and block jackknife standard error for cell type-specific open chromatin accounting for control peaks and baseline annotations. Summary statistics from the International Genetics of Ankylosing Spondylitis Consortium (IGAS) (**D**) and UK Biobank (**E**) GWAS were used. * indicates P < 0.05.

We validated this finding in a GWAS with genome-wide genotyping using the summary statistics for AS from the UK Biobank. With this GWAS, we confirmed that open chromatin regions specific for NK cells were significantly enriched in AS heritability (P = 0.034, **Fig. 1E**). To evaluate the reliability of our results, we included four control traits that have been extensively examined in similar studies integrating GWAS with functional genomics (15,18–20). As expected, RA presented the highest enrichment for T cell-specific open chromatin regions (P = 0.0018), Alzheimer’s for myeloid DC (P = 0.00018), and SLE for B cells (P = 0.0015). We selected body height as a negative control trait anticipating no significant enrichment for immune cells, a prediction that was confirmed by our data (all P > 0.1, **Supplementary Fig. 1**). Collectively, these epigenomic analyses suggest that AS risk alleles are preferentially located in regions that may influence gene regulation in NK cells.

To corroborate these findings using an alternative experimental approach, we used our previously published RNA-seq dataset of sorted peripheral CD4+ T, CD8+ T, MAIT, invariant NKT (iNKT), γδ T cells expressing Vδ1 TCR chain (Vd1), γδ T cells expressing Vδ2 TCR chain (Vd2), and NK cells (each in duplicate from 6 healthy donors, **Fig. 2A**) (28). We applied the SNPsea method, which quantifies enrichment of cell type-specific gene expression in risk loci for a given trait (conceptual scheme in **Fig. 2B**) by employing a non-parametric statistical method to calculate empirical P-values through comparison with sets of null SNPs (29). We used the AS risk SNPs reported by Brown and Wordsworth in 2017 which were curated from multiple AS genetic studies (30). SNPsea analysis revealed a significant enrichment of NK cell-specific gene expression in AS risk loci (P = 0.01), which was not observed in the other lymphocyte subsets included in the dataset (**Fig. 2C**).

**Figure 2.**
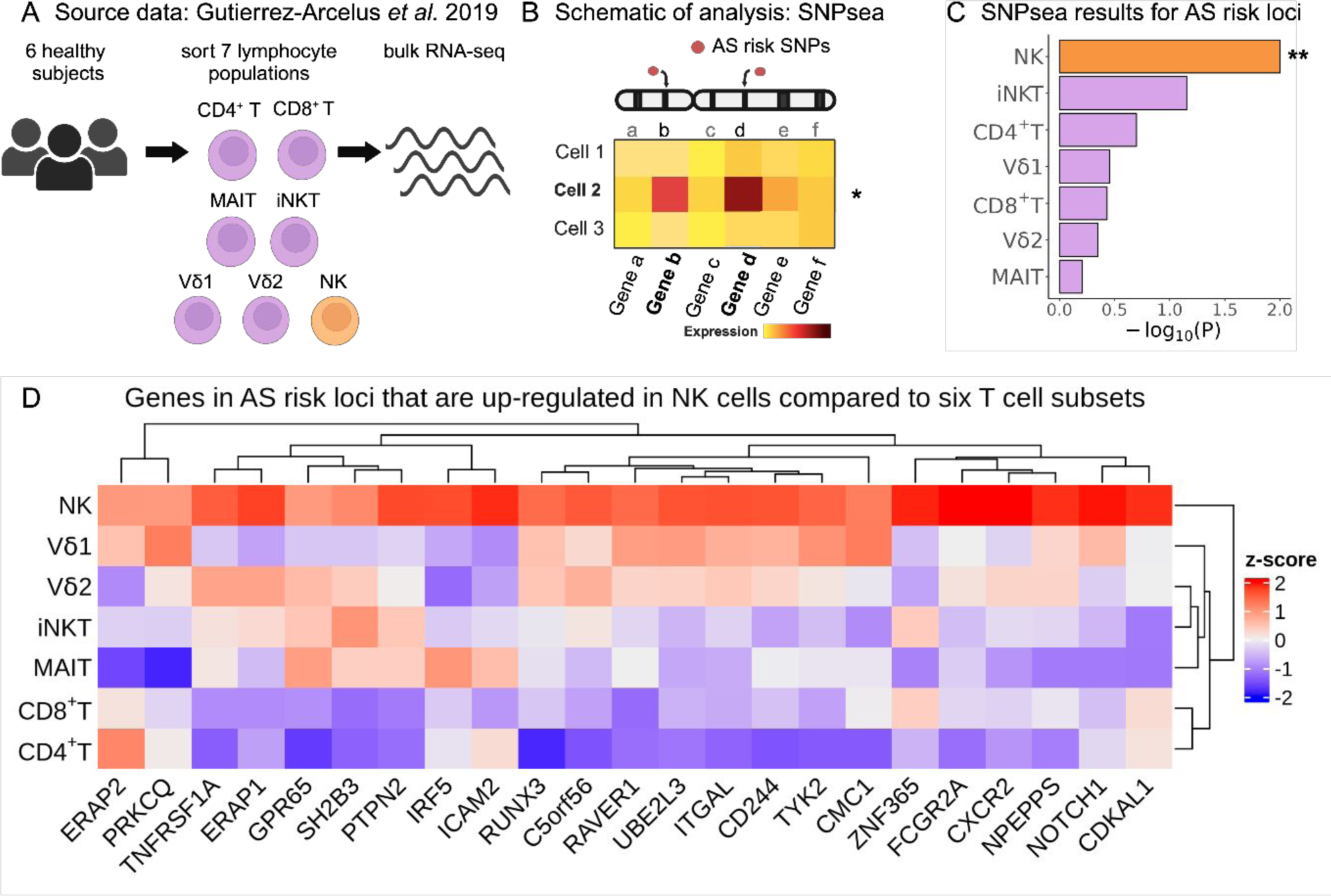
NK cells show enrichment of cell type-specific expression of AS-associated genes. (**A**) Cartoon depicting the Gutierrez-Arcelus *et al.* study. Peripheral blood cells from 6 healthy subjects were sorted into NK cells (orange) and six T cell populations (purple): CD4+ T, CD8+ T, MAIT, iNKT, and two γδ T cell populations. Bulk RNA sequencing was performed on two replicates per sample. (**B**) Graphical representation of the SNPsea method illustrating the integration of gene expression profiles with risk loci obtained from GWAS. (**C**) Bar graphs showing -log10(P-value) for enrichment of cell type-specific expression of genes in AS risk loci using SNPsea. (**D**) Heatmap showing expression levels for genes in AS risk loci that were significantly upregulated in NK cells compared to six T cells subsets. Expression levels are scaled by row. Tpm: transcripts per million. ** indicates P < 0.01.

We then performed a differential expression analysis comparing NK cells with the six T cell subsets (Supplementary Table 1). Genes in AS risk loci with significant upregulation in NK cells are presented in **Fig 2D**. Two of these genes, *RUNX3* and *TBX21* (Tbet), encode transcription factors with important roles in lymphocytes. *TNFRSF1A* encoding TNF Receptor I has a well-established association with AS that has been validated by multiple studies (31–33). *FCGR2A* codes for the low-affinity Fcγ receptor IIA, an activating receptor involved in orchestrating immune response. Less studied genes included *NPEPPS,* which encodes a puromycin-sensitive aminopeptidase, and *LNPEP*, which encodes a zinc-dependent aminopeptidase (34). Both genes are paralogs of *ERAP1* and belong to the MHC Class I antigen processing and presentation pathway, along with other known AS risk genes (35–37). Collectively, the results of our second integrative analysis indicate that several genes within AS risk loci are highly expressed in NK cells relative to T cells, providing additional support for the emerging hypothesis that AS risk alleles exert their effects, at least in part, via NK cells.

The transcriptomic phenotype of immune cells commonly differs between blood and tissue (38–41). Hence, in addition to analyzing peripheral blood as in the previous analyses, we sought to evaluate disease-relevant cell subsets from a tissue relevant for AS. We used the human Space-Time Gut Cell Atlas (42) which includes scRNA-seq data for samples from various locations of fetal (N = 16), pediatric (N = 8) and adult (N = 13, including 6 healthy and 7 Crohn’s disease patients) intestine (**Fig. 3A)**. We applied the scDRS method (16) which identifies cells that over-express a significant proportion of genes implicated by GWAS, weighted on their strength of association with disease, compared to null sets of control genes in the dataset (conceptual scheme in **Fig. 3B**). The Space-Time Gut Cell Atlas investigators identified the following broad cell types: mesenchymal, epithelial, endothelial, neuronal, myeloid, red blood cells, B cells, plasma cells, T cells, NK cells and other innate lymphoid cells (ILCs)( **Fig. 3C)**. scDRS identified 1,852 cells with significantly enriched expression of AS GWAS genes (20% FDR, **Fig. 3D**). Of these, 765 were T cells, 264 myeloid cells, 320 NK cells and 319 other ILCs. Normalized for cell type abundance in the dataset, NK cells showed the highest enrichment (39-fold), followed by other ILCs (34-fold), T cells (5-fold), and myeloid cells (5-fold, **Fig. 3E**). In contrast, non-immune cell types exhibited a depletion of disease relevant cells relative to their abundance in the entire dataset (**Supplementary Fig. 2**). We then used the fine-grained annotations of the Space-Time Gut Cell Atlas to identify the particular cell subsets that had significant expression enrichment of AS-associated genes. This revealed NK cells as the most abundant (N = 320), followed by Lti-like NCR+ ILC3 cells (N = 147), activated CD8+ T cells (N = 132), macrophages (N = 130), Lti-like NCR-ILC3 cells (N = 112), γδ T cells (N = 94), and other T cells, ILCs and myeloid subsets (**Fig. 3F**). Genes in AS risk loci with high expression in gut NK cells include *GNLY*, *CCL4* and *CCL3* (**Fig. 3G**). Using the control traits specified earlier, we confirmed T cells as the main disease-relevant cell type for RA and monocytes for Alzheimer’s disease (**Supplementary Fig. 2**). No significant disease-relevant cells were identified for height (as expected) and for SLE, which could mean that B cells in the gut are in a state not pertinent to SLE or that the dataset lacked sufficient power to detect an association for this disease (**Supplementary Fig. 2**). In sum, our analyses indicate that tissue-resident NK cells exhibit significant expression of AS-associated genes.

**Figure 3.**
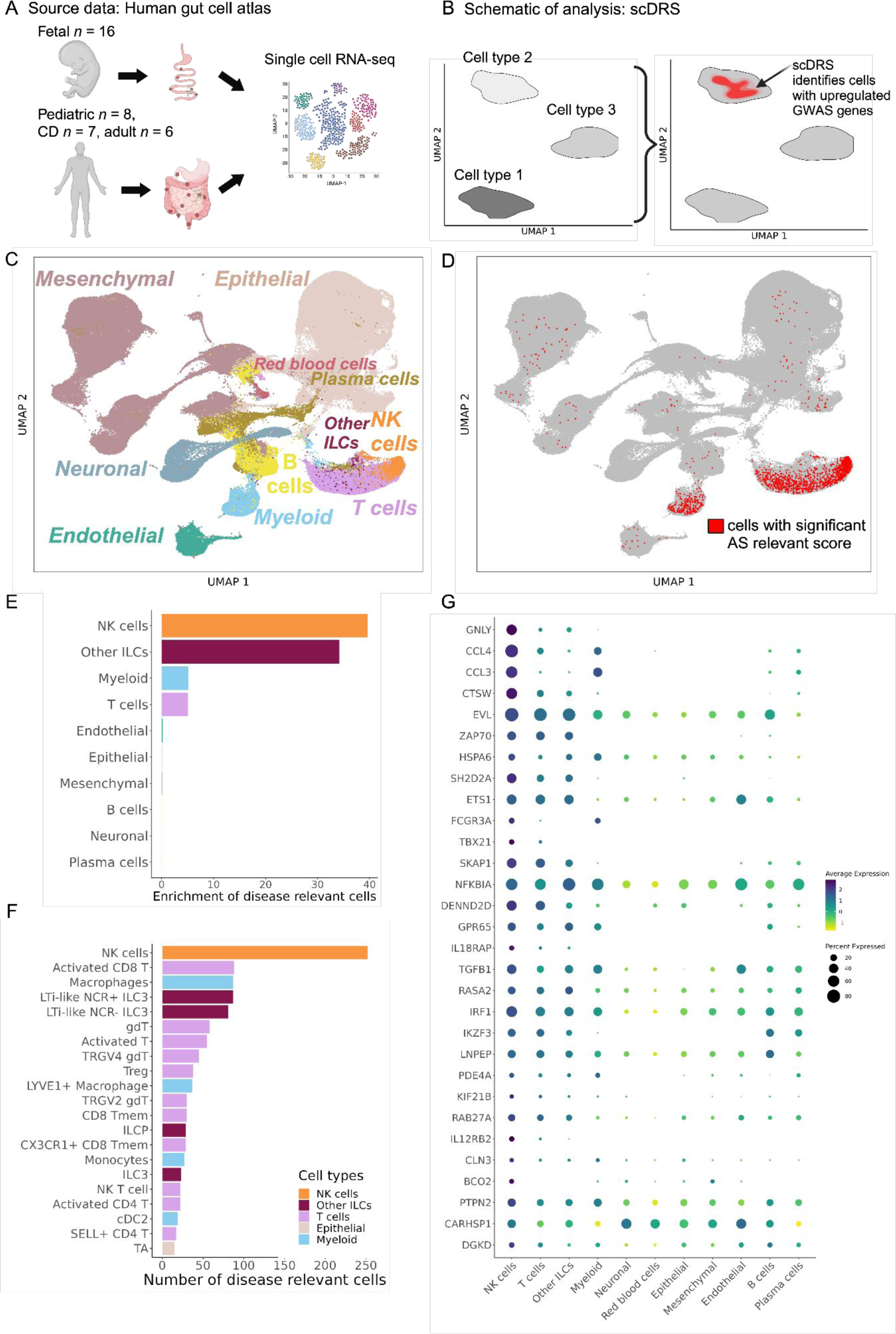
Human gut single-cell atlas reveals significant upregulation of AS-associated genes in NK cells. (**A**) Cartoon depicting the generation of the Space-Time Gut Cell Atlas with samples from fetal, pediatric and adult subjects. (**B**) Graphical representation of the scDRS method, which integrates GWAS risk genes with single cell data to identify disease-relevant cells. (**C**) Visualization of the Space-Time Gut Cell Atlas data using UMAP on the top 20 principal components from 1,997 variable genes from the scRNA-seq expression matrix. (**D**) Same UMAP visualization as in C. Cells with significant scDRS score (20% FDR) are colored in red. (**E**) Bar graph showing enrichment of scDRS significant cells per cell type (cell-type percent in whole dataset over cell-type percent within scDRS significant cells). (**F**) Bar graph showing the number of significant scDRS cells for each cell type using the fine-grained annotations from the Space-Time Gut Cell Atlas. Cell populations with at least 15 significant scDRS cells are shown. (**G**) Scaled average expression levels and percent of cells expressing a given gene for 50 genes associated with AS that had significant upregulation (5% FDR) in NK cells compared to the other cell types. Genes are sorted by multiplying their MAGMA score (strength of association with AS) by their average level of expression in NK cells.

Lastly, we sought to find putative target genes for AS risk variants in NK cells. To this end we performed co-localization analyses between AS GWAS risk loci and genetic variants associated with gene expression (expression quantitative trait loci, eQTLs) using coloc (43). We leveraged eQTL summary statistics from the eQTL Catalogue (44) drawing upon data from a study on the transcriptomic profiling of peripheral NK cells from 91 genotyped individuals (45) as well as a microarray QTL study that profiled NK cells from 245 genotyped individuals (46). We found four AS risk loci with genome-wide significance (P < 5 × 10^-8^) and a high posterior probability (>0.8) of sharing a causal variant with an NK cell eQTL (PP4, **Table 1**, **Fig. 4**). An additional 10 loci with suggestive AS association P-values (3.56 × 10^-5^ < P < 5.40 x 10^-8^) showed evidence of co-localization with NK cell eQTLs for 18 genes (PP4 > 0.75, **Table 1**). Within the genome-wide significant loci we identified the established target genes *ERAP1* and *TNFRSF1A*, as well as the putative target genes *ENTR1* (a.k.a. *SDCCAG3*) and *B3GNT2*, which have been studied less.

**Figure. 4.**
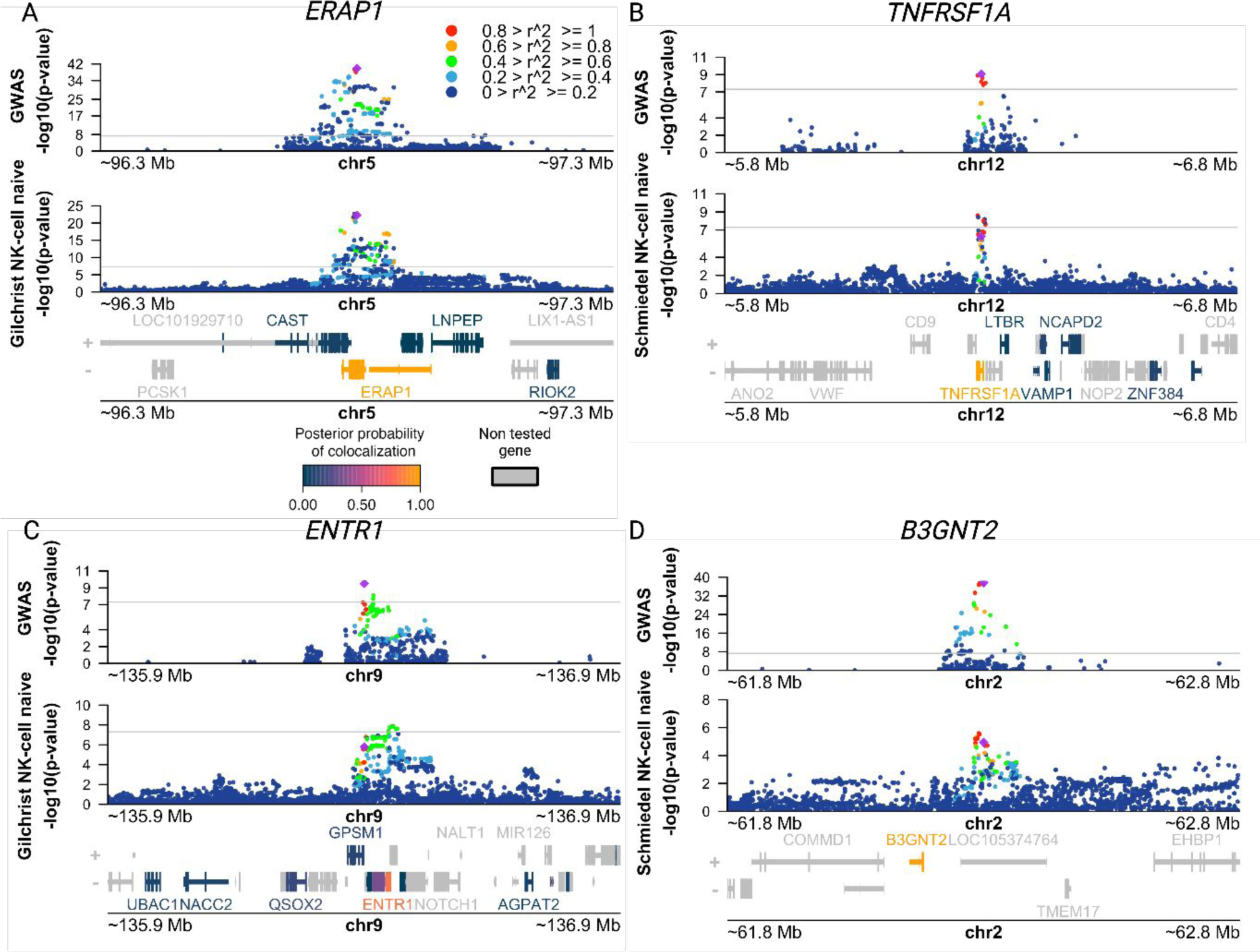
Co-localization of AS risk loci and NK cell eQTLs points to putative target genes for AS risk variants. (**A-D**) Manhattan plots showing AS GWAS and NK cell eQTL -log10(P-values) for SNPs within 500 kb of a lead GWAS SNP. The color of each SNP indicates its level of linkage disequilibrium (LD) between with the lead GWAS SNP (purple diamond). Genes in the region are colored according to their posterior probability of hypothesis four (PP4), i.e. that the same causal variant is shared between AS and the eQTL for that gene. (**A**) Manhattan plots identifying putative target gene *ERAP1* using AS IGAS GWAS (top) and NK microarray gene expression QTL (eQTL) data obtained from Gilchrist *et al*. (bottom) (**B**) Manhattan plots identifying putative target gene *TNFRSF1A* using AS IGAS GWAS (top) and NK gene expression QTL (eQTL) data obtained from Schmiedel *et al*. (bottom) (**C**) Manhattan plots identifying putative target gene *ENTR1* using AS IGAS GWAS (top) and NK microarray gene expression QTL (eQTL) data obtained from Gilchrist *et al*. (bottom) (**D**) Manhattan plots identifying putative target gene *B3GNT2* using AS IGAS GWAS (top) and NK gene expression QTL (eQTL) data obtained from Schmiedel *et al*. (bottom). All QTL summary statistics taken from eQTL Catalogue.

**Table 1.**
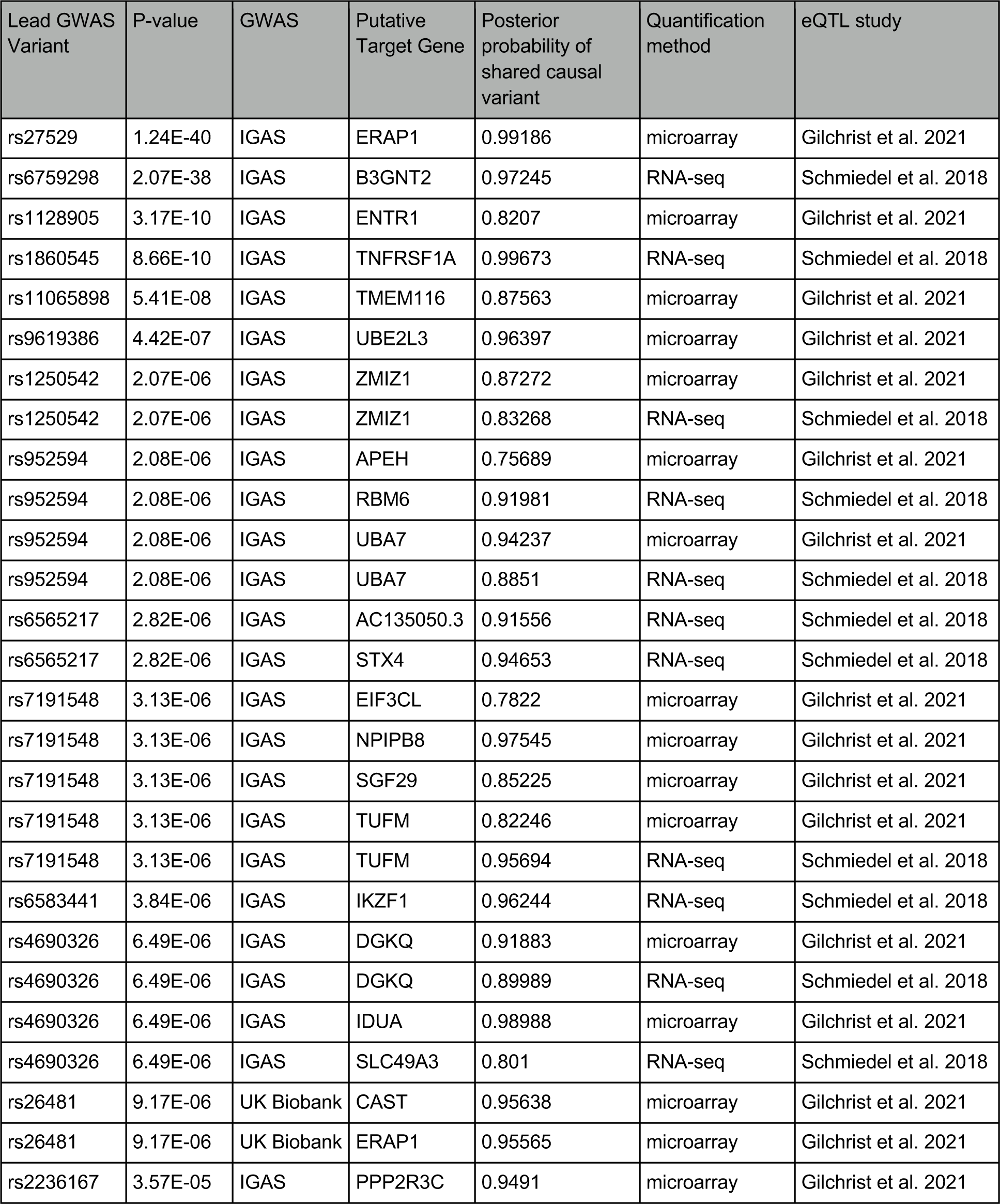
Putative target genes identified by co-localization analysis between AS-associated loci and eQTLs in NK cells.

## Discussion

In this study we integrated epigenomic and transcriptomic datasets with AS genetic risk data to find candidate cellular drivers of AS pathogenesis. Our unbiased approach, applying three different methods to datasets from both peripheral blood and tissue, consistently identified NK cells as the dominant disease-relevant cell type. Specifically, we found that NK-specific open chromatin regions and NK-specific gene expression were significantly enriched for non-MHC AS genetic risk. This suggests that a significant portion of AS risk variants affect gene regulation in NK cells, pointing to NK cells as potential key mediators of AS pathogenesis.

NK cells have the ability to directly destroy target cells through cell lysis, and in addition play a significant role in shaping immune responses by releasing cytokines (47). Previous studies support a role for NK cells in AS. AS patients with chronic subclinical intestinal inflammation were found to have an increased abundance of NKp44+ NK cells in their gut, and these cells were the major producers of IL-22 in the lamina propria, suggesting a possible role in tissue protection (48). One could speculate that dysfunctional NK cells “drive” AS development by contributing to intestinal inflammation, in line with the gut-joint axis hypothesis (49). Alternatively, NK cells may play a critical role through activities in spinal tissues. Cuthbert *et al.* studied entheseal immunology using discarded surgical specimens from patients with back pain (not axSpA) undergoing laminectomy and reported that NK cells are present in both entheseal soft tissue and peri-entheseal bone (50). We are not aware of any data assessing the presence of NK cells at spinal enthesis in AS patients or in the subchondral bone marrow in patients with sacroiliitis.

HLA-B27 can bind to the killer cell immunoglobulin-like receptor (KIR) KIR3DL1 and affect the function of NK cells, including their ability to lyse cells (51–53). HLA-B27 homodimers can also bind KIR3DL2 (54). Chan and colleagues showed an expansion of KIR3DL2+ NK and CD4+ T cells in AS patients (Chan Arthritis Rheum 2005, PMID 16255049). Subsequent studies by the same group focused on CD4+ T cells demonstrating that KIR3DL2+ CD4+ T cells were major IL-17A producers (55). However, an expansion of KIR3DL2+ NK and T cells has not been observed in other axSpA cohorts (56,57). Multiple risk loci for AS include genes relevant for NK cell function, including *KIR2DL1*, *KIR3DL1*, *KIR2DS5*, *KIR3DS1* and *KIR2DL5* (58–62). In another study, investigators co-cultured ERAP1-inhibited M1 macrophages with NK cells from AS patients, and found that patients with ERAP1 protective alleles led to decreased CD69 and CD107a on NK cells and a lower number of IFN-γ+ NK cells compared to patients carrying non-protective alleles (63).

Our findings do not rule out the involvement of other cell types in AS pathogenesis. Indeed, in the human Space-Time Gut Cell Atlas, we identified significant expression of AS-associated genes in T cell subsets and ILC subsets (Fig. 3D-F), which share transcriptional programs with NK cells (28,64–66). Indeed, it is likely that genetic risk to AS is mediated through multiple cell types, as is the case for other complex diseases such as multiple sclerosis, for which studies have found risk enrichment in open/active chromatin regions specific to both T cells and B cells (20). We and others have shown that eQTLs often exhibit impact in multiple cell types (67,68). Hence, determining the specific cell type through which a disease risk variant is exerting its pathogenic effects can be challenging.

Our co-localization analyses using two eQTL NK cell datasets identified four putative target genes for AS risk variants: *ERAP1*, *TNFRSF1A*, *ENTR1* (a.k.a. *SDCCAG3*) and *B3GNT2*. The importance of *ERAP1* in AS risk is well established, and polymorphisms affecting its expression have been reported for multiple cell types, including macrophages, monocytes, T cells, and induced pluripotent stem cells, fibroblasts, and immortalized B cells (69–71). Similarly, multiple studies have found significant associations between non-coding polymorphisms at or near *TNFRSF1A* and AS, including in European and East Asian populations (31,32,72,73). While there are multiple genes in this genomic locus, including *PLEKHG6*, *SCNN1A*, and *LTBR*, our co-localization results suggest that *TNFRSF1A*, which encodes TNF receptor I, is the target gene of the causal variant in this locus, and its dysregulation can happen in NK cells. This is consistent with the therapeutic efficacy of TNF inhibitors in AS and the known function of TNF as a booster of the cytolytic capacity of NK cells (74). Interestingly, TNFRSF1A has been functionally linked to ENTR1, a less extensively studied putative target gene identified in this study. *ENTR1*, which encodes an endosome-associated trafficking regulator, is needed for TNF receptor expression on the cell surface (75). Lastly, *B3GNT2* encodes an acetylglucosaminyltransferase enzyme that is a type II transmembrane protein. A recent study in a Taiwanese cohort demonstrated that a non-coding genetic variant near *B3GNT2* is associated with AS susceptibility, and that *B3GNT2* blood mRNA levels were negatively correlated with C-reactive protein (CRP), erythrocyte sedimentation rate, syndesmophyte formation, and Bath ankylosing spondylitis functional index (BASFI) (76).

While the 4 putative target genes identified here make sense in the context of AS and potential impact on NK cell function, they are not as numerous as we would have expected given the 32 genome-wide significant risk loci included in the co-localization analyses. However, similar challenges have been encountered with non-coding risk variants in other complex diseases, where only 20-47% of risk variants co-localized with eQTLs (77–79). Our research, along with that of others, suggests that many regulatory effects might remain undetected due to their presence in cell states of activation or differentiation that have not been thoroughly explored (80–83). Moreover, the sample size of typical eQTL studies is likely insufficient to find the regulatory effects of most risk variants identified by GWAS (84). Hence, we believe that better-powered eQTL studies ascertaining multiple activation states in NK cells are needed to find additional target genes for AS risk variants.

One limitation of our study, due to a lack of published data, is the incomplete assessment of the spectrum of potentially relevant immune cell subsets and states, particularly those present in inflamed sacroiliac joints and spine. Consequently, if the real driver for AS pathogenesis is a cell subset or state that was not present in the analyzed datasets, but has transcriptomic and epigenomic similarities to NK cells, then our results may suffer from “guilt-by-association” bias. To our knowledge, current transcriptomic datasets profiling multiple immune cell types from AS patients are limited to peripheral blood (14,85–90). When we applied scDRS to a recently published single-cell RNA-seq dataset of 98,884 PBMCs cells from 10 AS patients and 29 healthy controls (88), we found no significant cells for the disease-relevant gene expression score (data not shown), possibly due to lack of power in the study for this type of analysis.

Our study encompassed a broad spectrum of immune cell states within the gastrointestinal tract and peripheral blood of healthy subjects and consistently pointed to NK cells. Since GWAS pinpoint genetic regions implicated in the onset of disease, including early stages when future patients are still asymptomatic, the study of samples from healthy subjects is relevant, despite the possibility that not all cell states are represented. Future investigations, particularly larger-scale studies of samples from blood and inflamed tissue from AS patients including untreated patients in the early phases of the disease, will be key to establish whether NK cells are indeed drivers of AS pathogenesis.

## Materials and Methods

### Genome-wide association studies

We used the GWAS ImmunoChip summary statistics from the International Genetics of Ankylosing Spondylitis Consortium (IGAS). The IGAS study, led by (7), performed high-density genotyping of 9,069 AS cases and 13,578 healthy controls. In addition, we used the GWAS summary statistics from the UK Biobank, which involved a case-control design with 1,185 AS cases and 419,276 controls, providing genome-wide coverage for AS susceptibility loci (91).

We lifted the genomic positions of the genetic variants to genome build hg19 or hg38 according to the version compatible with subsequent analyses. Given the complexity and strong genetic association signals within the Major Histocompatibility Complex (MHC) region, we excluded variants located on chr6:25,000,000–34,000,000.

We additionally used GWAS summary statistics for rheumatoid arthritis, Alzheimer’s disease, and systemic lupus erythematosus as positive control traits for which we know the disease relevant immune cell types, and height as a negative control trait for which we do not expect immune cells to be relevant. The summary statistics for control traits were preprocessed by the Alkes Price laboratory, they included HapMap 3 SNPs and SNPs that are in the 1000 Genomes Project, and they excluded the MHC region (chr6:25Mb-34Mb). These summary statistics are available at https://alkesgroup.broadinstitute.org/.

### Epigenomic and transcriptomic datasets

To identify cell type-specific open chromatin regions in different immune cell types, we used the Calderon *et al.* study (*19*), in which the authors collected blood from 4 healthy subjects, sorted immune cell types, and generated chromatin accessibility profiles using Assay for Transposase-Accessible Chromatin sequencing (ATAC-seq, GSE118189).

To find AS risk enrichment for cell type-specific expression, we incorporated data from the study conducted by Gutierrez-Arcelus *et al.* (28), which involved low-input mRNA-seq data from sorted NK cells and six T cell subsets isolated from 6 healthy subjects (each with two replicates per cell-type, GSE124731).

We used the Space-Time Gut Cell Atlas to identify cells exhibiting significant upregulation of disease-associated genes. This dataset includes single-cell RNA-seq profiling of 428,000 intestinal cells obtained from fetal (N = 16), pediatric (N = 8), and adult donors (N = 13). The dataset covers 11 different intestinal regions (42), https://www.gutcellatlas.org/.

### Differential accessibility analysis

We used the counts of open chromatin consensus peaks called by Calderon et al. First, we transformed counts into Reads Per Kilobase per Million mapped reads (RPKM), then normalized by quantiles using the preprocess Core R package and finally scaled to their log2 (normalized RPKM+1), thus we account for differences in library size across samples and peak length variability. We pooled sorted samples into 7 main immune cell types, aiming for a similar number of samples per cell type to avoid biases in the differential accessibility analyses: T cells (stimulated and unstimulated CD8+ T, unstimulated naïve CD4 T and memory CD4 T), B cells (stimulated and unstimulated bulk B cells, unstimulated memory and naïve B cells), natural killer cells (stimulated and unstimulated mature NKs, unstimulated memory NK and immature NK cells), monocytes (stimulated and unstimulated monocytes), plasma cells (unstimulated plasma cells), dendritic cells (unstimulated myeloid DCs), plasma dendritic cells (unstimulated plasmacytoid DCs). The latter three cell-types had less samples available, however, this did not impede our control trait Alzheimer disease to show significant heritability enrichment for myeloid DC-specific open chromatin regions, as expected (see Methods below and main text).

Next, we employed linear mixed model regression to identify regions that exhibited differential accessibility between each cell type and the rest of the cell types. To account for potential donor-specific effects, we incorporated the donor ID variable as a random effect in our analysis. For each cell type comparison, we tested peaks that had counts greater than the mean for that cell type in at least half of the samples, this yielded between 400 to 600 thousand tested peaks depending on the cell type. To select the “cell type-specific open chromatin peaks” for each cell type, we sorted open chromatin peaks by their t-statistic and chose the positive top 10%.

### Partitioned heritability enrichment analysis with LDSC-SEG

Linkage Disequilibrium score regression applied to specifically expressed genes (LDSC-SEG) v1.0.1 method was applied to determine disease-relevant cell types (15,92) for AS. Cell type-specific open chromatin peaks were extended by 225bp to each side, to match the genomic coverage recommended by the LDSC-SEG authors. These annotations were then utilized as input for the partitioned heritability enrichment analysis by LDSC-SEG. We used the baseline annotation v1.2 provided by Price Lab for LDSC-SEG, comprising 75 background annotations. Additionally, we used all consensus peaks (N = 829,942) of Calderon et al. as the control annotation. Using other baselines or controls did not affect our results. We utilized SNP weight files derived from the HapMap 3 project (HM3) European population.

### Analysis of cell type-specific gene expression enrichment in risk loci using SNPsea

SNPsea analysis aimed to assess the association between risk SNPs and genes expressed specifically for a given cell type (29). We incorporated a curated list of risk SNPs for ankylosing spondylitis (AS), compiled by Brown and Wordsowrth 2017 (30), which includes genetic variants that have been associated with AS susceptibility. This list was derived from multiple AS studies conducted until 2017.

We utilized the expression data obtained from Gutierrez-Arcelus et al. (2019). The gene expression counts in this dataset were normalized to transcripts per million (TPM) and transformed to log2(TPM+1) values. To identify the genes with meaningful expression levels, we included those with log2(TPM+1) > 2 in at least 10 samples. SNPsea was then run for the normalized expression matrix and AS risk SNPs, using recombination intervals from Myers et al. (93), null SNPs from Lango et al. (94), and the following parameters: --score single --slop 10000 --threads 2 --null-snpsets 0 --min-observations 100 --max-iterations 10000000.

### Integration of GWAS with single-cell RNA-seq with scDRS

We used the single-cell Disease Relevance Score (scDRS) by combining scRNA-seq and GWAS to identify cells with significant up-regulation of disease-associated genes, which are scored based on their strength of association with disease, and are compared with null sets of genes present in the dataset.

As recommended by scDRS authors, we first created disease-relevant genesets using Multi-marker Analysis of GenoMic Annotation (MAGMA) version 1.10 (95). First, we generated gene annotations with MAGMA setting a window of 10kb using the following parameters: “--annotate window=10,10 --snp-loc ./g1000_eur/g1000_eur.bim --gene-loc ./NCBI37.3/NCBI37.3.gene.loc”. Then we ran MAGMA using GWAS summary statistics for traits of interest with the following parameters: --bfile ./magma_v1.10/g1000_eur/g1000_eur --pval GWAS.pval use=’SNP,P’ ncol=’N’ --gene-annot ./magma_v1.10/out/step1.genes.annot.

We ran scDRS using the disease-relevant gene sets from MAGMA, the expression data obtained from the Space-Time Gut Cell Atlas (42) and corrected for biases by adding as covariates the number of genes expressed per cell and sample batch. Next, for visualization purposes and downstream analysis, we processed the single-cell dataset using Seurat (96), we performed integration across batches with Harmony (97), and we visualized cells in two dimensions with Uniform Manifold Approximation and Projection (UMAP). We labeled cells plotted in UMAP by the annotations defined by the Space-Time Gut Cell Atlas. Additionally, we colored cells by their scDRS score when cells passed the 0.20 FDR threshold.

### eQTL co-localization analysis

To select genomic loci for colocalization analysis, GWAS summary statistics were sorted by P-values, then starting from the variant with the smallest P-value, variants within a 50 Kb window were removed. The process was repeated with the next most significant variant among the remaining variants until no variant with a P-value below 5 x 10^-5^ was left. We performed colocalization analysis for GWAS studies against the eQTL Catalogue (44). We imported eQTL summary statistics from RNA-seq and microarray from Schmiedel et al. (45) and Gilchrist et al. (2022). We fetched the summary statistics data using the tabix method with the seqminer R package (v8.5). For each region tested, we included all biallelic SNPs that were ascertained in both the GWAS and eQTL study and performed the analysis only for genes within a window of ±500,000 base pairs from the GWAS top variant, and for which there was at least one eQTL passing the 5 x 10^-5^ P-value threshold. Before merging GWAS and QTL data, the variant coordinates of the GWAS were lifted to the GRCh38 version of the reference genome using liftOver with the UCSC chain file. We used the coloc v5.1.0.1 package (98) in R v4.1.0 to test for colocalization at each gene and dataset. The code is available at <https://github.com/gutierrez-arcelus-lab/>.

Each locus was plotted using plotgardener (99), and we recovered the LD of the top SNP in a given region in the GWAS dataset using the locuscomparer package (100). Then we used plotgardener functions to display the regions near the lead variant and colored the genes tested using the posterior probability that the two traits share a causal variant (PP4).

## Data availability

All data and methods are publicly available as specified above.

## Acknowledgments

This study was supported by a seed grant from the Spondyloarthritis Research and Treatment Network (SPARTAN) and a microgrant from the Joint Biology Consortium (1P30AR070253-01). PAN was supported by P30AR070253 and R01AR073201. JE was supported by NIH Grant R21 AR076040-01 and an ASPIRE grant from Pfizer. MGA was supported by P30AR070253, the Arthritis National Research Foundation, the Lupus Research Alliance, and the Gilead Sciences Rheumatology Research Scholars Award. We thank Soumya Raychaudhuri and Kamil Slowikowski for guidance on implementing SNPsea, and Steven Gazal for guidance on implementing LDSC-SEG. We thank the Gutierrez-Arcelus and Nigrovic laboratories for feedback on this study.

---

**Supplementary Figure 1.**
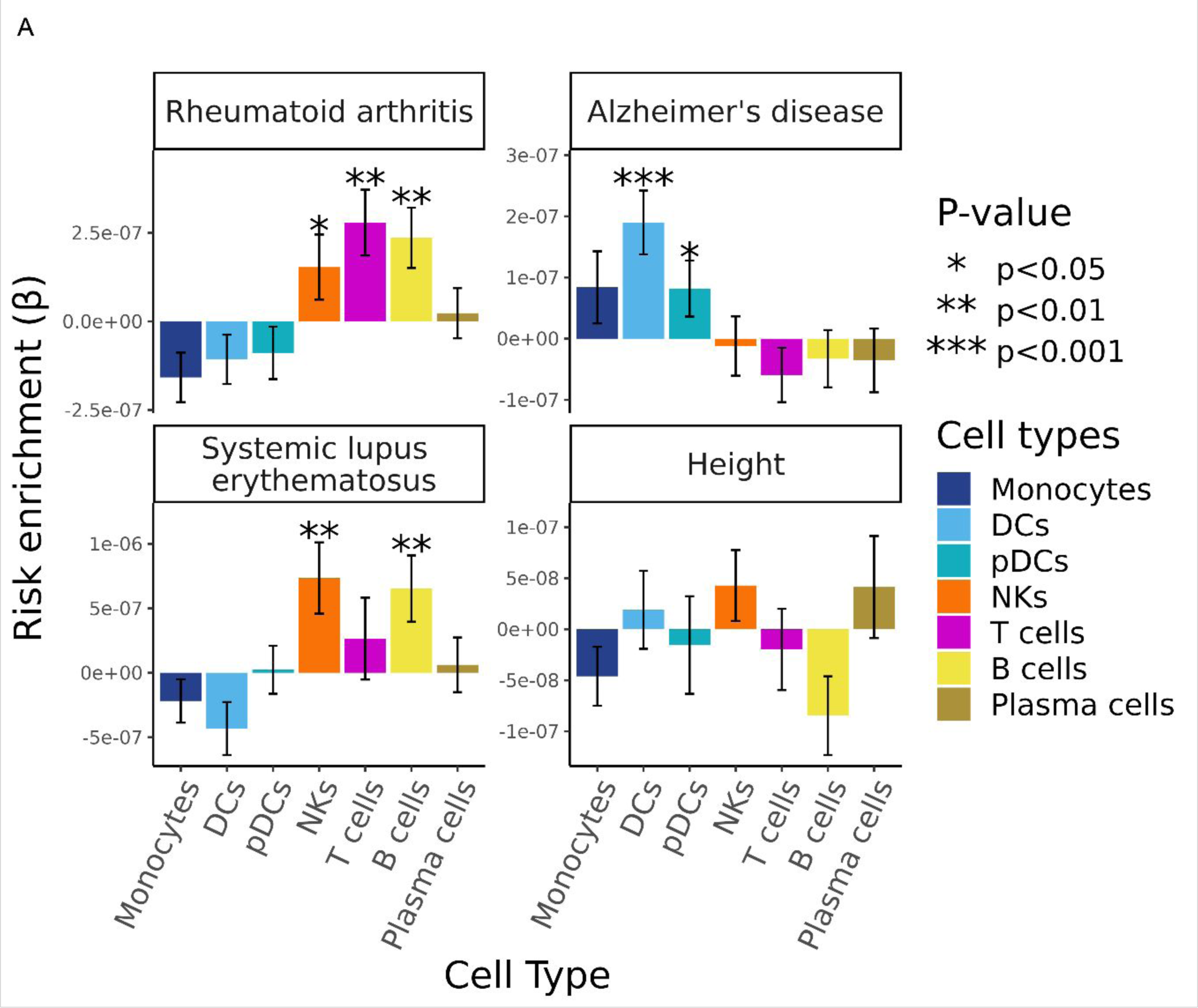
Heritability enrichment results for control traits. (**A**) Bar graphs display the genetic risk enrichment coefficient (y-axis) and standard error for cell-type specific open chromatin accounting for control peaks and baseline annotations. Open chromatin data were taken from the Calderon *et al*. study. Risk enrichment was assessed using GWAS summary statistics for the positive control traits rheumatoid arthritis, Alzheimer’s disease, systemic lupus erythematosus, and the negative control trait height. Bars marked with “*” indicate P < 0.05, “**” indicates P < 0.01, “***” indicates P < 0.001.

**Supplementary Figure 2.**
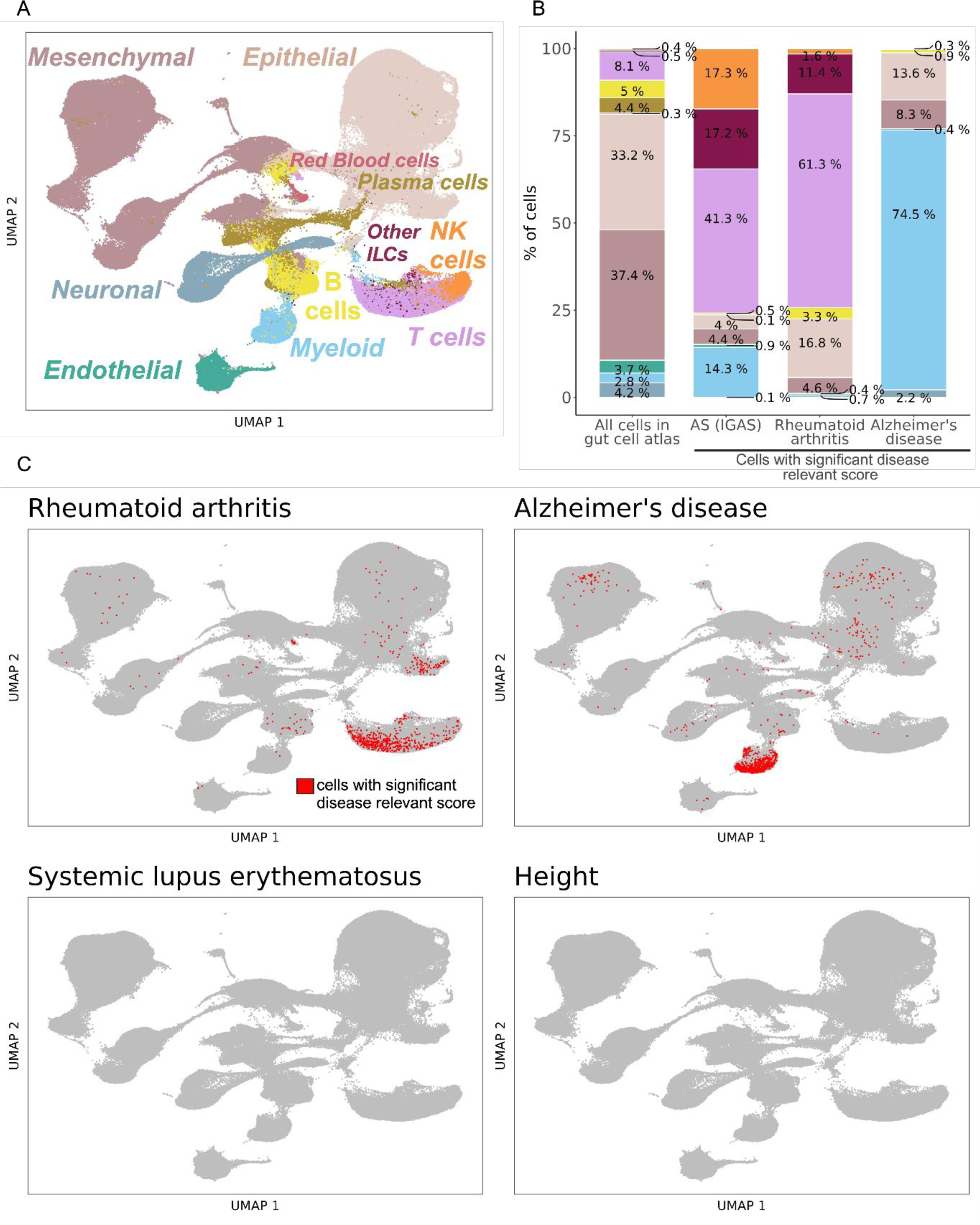
Single-cell disease relevant score results for control traits. (**A**) Visualization of the Space-Time Gut Cell Atlas using Uniform Manifold Approximation and Projection (UMAP) on the top 20 principal components from 1,997 variable genes from the single-cell RNA-seq expression matrix. Cells are colored based on the coarse cell type annotations from the Space-Time Gut Cell Atlas. (**B**) Barplots shows the cell type proportions within the whole Space-Time Gut Cell Atlas and within cells with significant disease relevant score (20% FDR) for AS (using IGAS GWAS), Alzheimer’s disease (AD) and rheumatoid arthritis (RA). (**C**) Same UMAP visualization as in A, where cells with significant scDRS score (20% FDR) are colored in red and non-significant cells are colored in gray, for each control trait.

